# Data Missingness in Digital Phenotyping: Implications for Clinical Inference and Decision-Making

**DOI:** 10.1101/2024.10.03.24314808

**Authors:** Joanna Shen, Kareem Abdelkader, Zacharia Isaac, Danielle Sarno, Jennifer Kurz, David Silbersweig, Jukka-Pekka Onnela, Daniel Barron

## Abstract

**Background:** Digital phenotyping, the use of personal digital devices to capture and categorize real-world behavioral and physiological data, holds great potential for complementing traditional clinical assessments. However, missing data remains a critical challenge in this field, especially in longitudinal studies where missingness might obscure clinically relevant insights.

**Objective:** This paper examines the impact of data missingness on digital phenotyping clinical research, proposes a framework for reporting and accounting for data missingness, and explores its implications for clinical inference and decision-making.

**Methods:** We analyzed digital phenotyping data from a study involving 85 patients with chronic musculoskeletal pain, focusing on active (PROMIS-29 survey responses) and passive (accelerometer and GPS measures) data collected via the Beiwe Research Platform. We assessed data completeness and missingness at different timescales (day, hour, and minute levels), examined the relationship between data missingness and accelerometer measures and imputed GPS summary statistics, and studied the stability of regression models across varying levels of data missingness. We further investigated the association between functional status and day-level data missingness in PROMIS-29 subscores.

**Results:** Data completeness showed substantial variability across timescales. Accelerometer-based cadence and imputed GPS-based home time and number of significant locations were generally robust to varying levels of data missingness. However, the stability of regression models was affected at higher thresholds (40% for cadence and 60% for home time). We also identified patterns wherein data missingness was associated with functional status.

**Conclusion:** Data missingness in clinical digital phenotyping studies impacts individual- and group-level analyses. Given these results, we recommend that studies account for and report data at multiple timescales (we recommend day, hour, and minute-level where possible), depending on the clinical goals of data collection. We propose a modified framework for categorizing missingness mechanisms in digital phenotyping, emphasizing the need for clinically relevant reporting and interpretation of missing data. Our framework highlights the importance of integrating clinical with statistical expertise, specifically to ensure that imputing missing data does not obscure but helps capture clinically meaningful changes in functional status.

## Introduction

Digital phenotyping has emerged as a promising tool for studying health and disease.^1–3^ Often defined as the use of personal digital devices like smartphones and wearables to capture “moment-by-moment,” real-world behavioral and physiological data, digital phenotypes offer a unique window into how individuals function in their daily lives.^4^ In particular, digital phenotypes can complement traditional clinical assessments and patient reported outcome measures (PROMs) which are often “intermittent, brief, and superficial cross-sectional examinations.”^5,6^ For example, in older patients with chronic pain conditions, we recently introduced High-frequency Ecological Recordings of Mobility, Emotion and Sociability (the HERMES phenotype) as a digital phenotype “panel” that measures functional status; mobility, emotion, and sociability being the primary indices of functional status.^2,7^ Long-term, the hope is that the HERMES phenotype (and overall, the HERMES approach) might offer three things: first, measures of an individual patients’ baseline function (how active they are); second, the ability to detect *changes* from this baseline that might portend a functional decline; and third, data to guide patient-specific, just-in-time clinical decisions.^2,6,8^ Digital phenotypes therefore promise a unique window into clinically-relevant behavior and clinical decision.^2,9^ However, a significant challenge is the frequent occurrence of missing data.^10,11^

Missing data can be simply defined as the absence of data points or values in a data set. This definition implies an *expectation*; that is, data can only be missing if they were *expected* to be present in a data set. The study of missing data in statistics attempts to solve a very specific problem: although many statistical methods have been designed for data sets with no missing values, missing values are common in practice in the social and health sciences, especially in longitudinal studies.^12,13^ Today, various techniques can mitigate challenges imposed by missing data, thereby allowing the analyses to proceed.^14,15^ Which missing data technique one should deploy depends on *why one believes* those data to be missing.

*Why* data are missing—also referred to as the “mechanism of missingness”—is a complex topic that depends on what, how, and why data were measured at all.^16^ Data can be Missing By Design (MDB), meaning that there are periods of time or types of data that are deliberately not collected (e.g., in digital phenotyping studies GPS data is typically collected only intermittently to save battery life). Beyond MBD, the “Mechanisms of Missingness” are often grouped in 3 categories: missing at random (MAR), missing not at random (MNAR), missing completely at random (MCAR). First described by Rubin, these mechanisms reference nonresponse in social science surveys and censuses.^16^ That is, the goal was to allow researchers to proceed with group-level descriptive statistics.^16^

Missing data is a particular problem in group-level clinical research, especially in clinical trials. In 2010, at the request of the U.S.A. Food and Drug Administration, the (U.S.) National Academy of Sciences (NAS) convened a panel and published a report on “The Prevention and Treatment of Missing Data in Clinical Trials.”^17^ The NAS report framed missingness as primarily a problem of attrition (i.e., when patients discontinue study participation) noting that the reasons why a person leaves a study may be directly related to the clinical trial and must therefore be accounted for (e.g., a patient discontinued study treatment due to inefficacy or side effects).

Missingness, the NAS stated, “reduces the benefit provided by [clinical trial] randomization and introduces potential biases in comparison of the treatment groups.”^17^ Because individual studies deploy bespoke, “ad hoc and variable” approaches to account for data missingness, the report recommends specific, “more principled” approaches to clinical trial design and analysis in hopes of bringing a standardized solution to the problem.

At the individual level, missing data present a complex challenge, particularly when data collection aims to guide clinical inference and decision-making in individual patients. Although missing data is a recognized issue in digital phenotyping research^18^, how to account for missingness and understand its mechanisms in the context of clinical research remains unresolved in the literature.^19,20^ This ambiguity has led to inconsistency in how missing data are reported in research papers and, more critically, in how to adjust for missing data in clinical research. For example, a digital phenotyping study might report an association between a digital measure (e.g., step count) and a clinical outcome (e.g., pain interference) without adequately addressing the extent and reasons for missing data or the potential impact on the study’s conclusions (e.g., patients with high pain interference might not charge their digital device, leading to greater missing data and thereby lower daily step count). This oversight is understandable, given the lack of clarity, but it impedes the clinical application of digital phenotyping research.

In this paper, we closely examine digital phenotyping data and, based on our analysis, propose a framework, terminology, and timescale for reporting missing data in digital phenotyping clinical research. We introduce new terminology to describe the mechanisms of data missingness, linking them directly to the clinical goals of data collection. Additionally, we outline a method for updating clinical inferences based on the extent of missing data.

## Methods

We sought to understand how missing data impacted statistical results in digital phenotyping clinical research and how data missingness could affect clinical decisions. To do this, we explored data collected as part of our study of functional status of older patients with chronic musculoskeletal pain conditions.^21^

### Participant population

All participants were recruited from the Pain Intervention & Digital Research Program (Pain-IDR), based in Spaulding Rehabilitation Hospital at Charlestown, MA. Our IRB protocol was approved and monitored by the MassGeneralBrigham IRB. We computed age based on each person’s start date on the study. In total, we recruited 85 participants with missing demographic information from 1 participant. Of the other 84 participants, the mean age was 55.2 (standard deviation of 15.7) with an age range of 19-84 years (see Table 1 for demographic information). In this sample, we had 69 iPhone and 15 Android users.

**Table 1.** Participant demographic information. A more detailed description of recruitment location and study setting can be referenced in Fu (2024).

|  | Age |  |  | Race |  |  |  |  |  |
| --- | --- | --- | --- | --- | --- | --- | --- | --- | --- |
|  | Mean | Std Dev | Range (Min-Max) | American Indian or Alaska Native | White | Asian | Black or African American | Other / Unknown | Total |
| Female | 52.65 | 15.49 | 23-80 | 1 | 38 | 2 | 5 | 5 | 51 |
| Male | 59.47 | 15.61 | 19-84 | 0 | 27 | 0 | 2 | 3 | 32 |
| MTF | 46 | 0 | 46-46 | 0 | 1 | 0 | 0 | 0 | 1 |
| Total | 55.17 | 15.74 | 19-84 | 1 | 66 | 2 | 7 | 8 | 84 |

### Study design

We have previously reported ^2^ our study design, but provide a brief overview here for the benefit of the reader. On consent and entry to the study, participants were guided to install the Beiwe application on their personal smartphone. From here, we collected two types of data: active and passive. Active data included daily “microsurveys” from the PROMIS-29 and a daily pain score (0-10). We standardized PROMIS-29 question responses by adjusting the Likert scale responses to range from “1” to “5” with “1” indicating the response suggestive of greatest health and “5” indicating a response suggestive of the worst health. Standardizing questions allowed us to more easily define consistent subscores for cross-subscore comparison (detailed below). Passive data included accelerometer and GPS data with summary measures computed by Forest, a library of data analysis tools and methods (forest.beiwe.org).

As previously reported, our original IRB protocol did not have a monthly minimum completion requirement.^21^ After observing that some participants no longer responded to the micro-surveys a few weeks after being onboarded on the study, we realized we needed a structured, formal way to dismiss people from the study. We therefore amended our IRB protocol so that study participants with 1 month of data with less than 20 percent survey completion would be notified of this and asked to troubleshoot; participants with more than 2 consecutive months of data with less than 20 percent survey completion would be dismissed from study. It was at this time we began analyzing the extent of missing data in our sample.

### Determining participant withdrawal and engagement

For each participant, we determined withdrawal days and hours by analyzing the presence or absence of data over the 180-day study period. While similar to the below-described data missingness and data completeness measure, we computed the day and hour-level presence or absence of data to investigate the following: 1) how data completeness change at day and hour-level data across the duration of the study and 2) when each participant withdrew from the study at the day and hour-level (see Figure 1).

**Figure 1.**
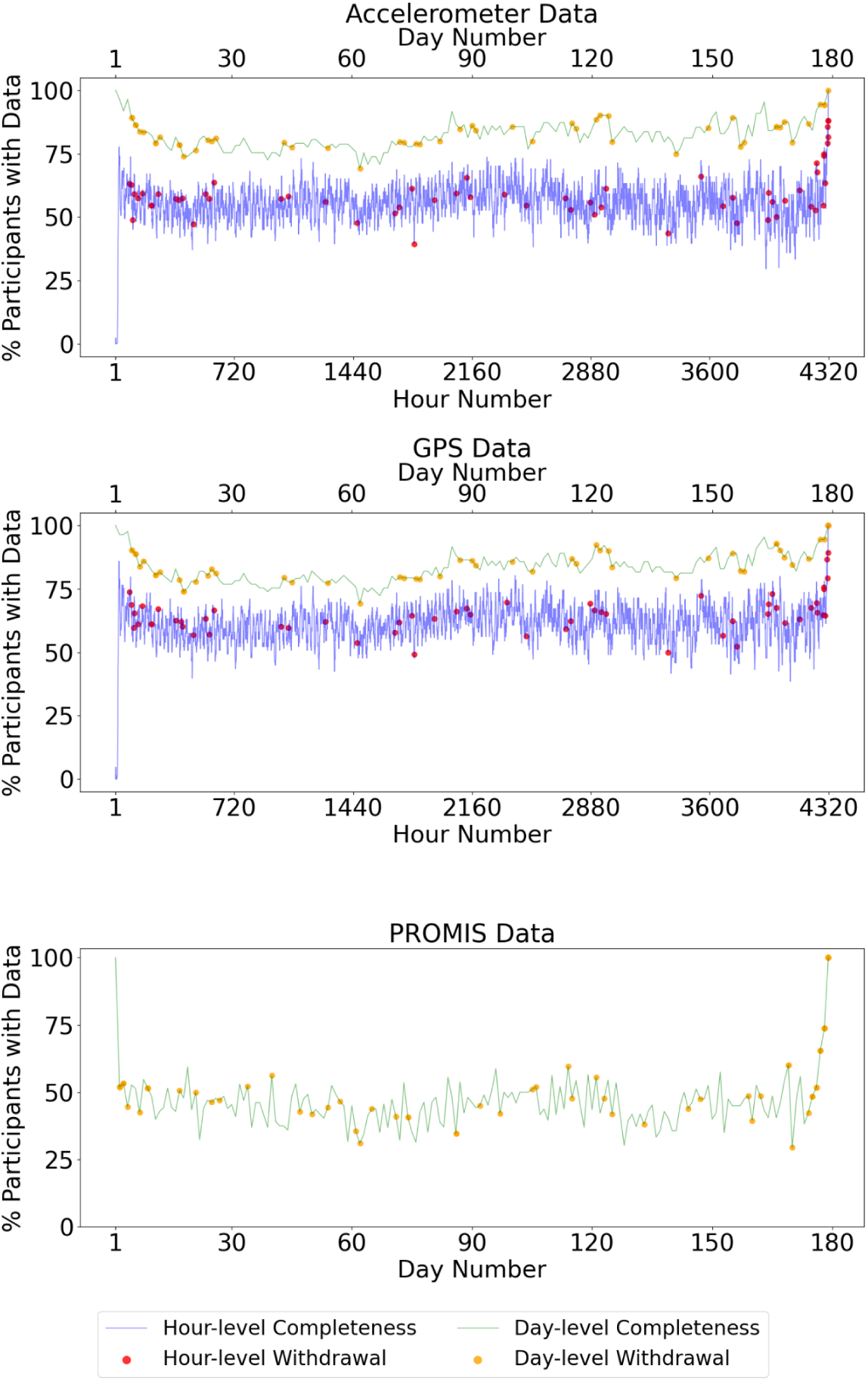
Data completeness trends over the 180-day study period. The figure shows day-level and hour-level data completeness for passive data (accelerometer and GPS) and active data (PROMIS-29) throughout the study. Day and hour numbers were standardized across participants based on their study initiation day 1 and hour 1. More withdrawals occurred during the first 30 days (or first 720 hours), as indicated by red and orange dots, leading to an early decline in completeness. However, completeness remains relatively stable mid-study, with an end-of-study artifactual increase due to fewer participants, which inflates the percentages. Passive and active data completeness often align; when active data is present, passive data would also be present, usually with a higher completeness rate.

To standardize the comparisons across participants, we aligned day and hour numbers based on each participant’s study initiation day and hour, with day 1 and hour 1 representing the start of their participation. This approach allowed us to systematically track when participants ceased providing data relative to their own study timelines. We assigned a binary indicator (1 for data present, 0 for data missing) at for each day and hour. Continuous data missingness from a specific time point onward was flagged as the participant’s withdrawal. We calculated the percentage of participants with data each day and hour, identifying trends in participant engagement over time. For active data (PROMIS data), unlike passive data (accelerometer and GPS data), only day-level withdrawal and engagement were studied, given that the survey prompts were administered once daily.

A participant was considered withdrawn from the study if they failed to provide any data – either passive or active – for the remainder of the study period, starting from the first day or hour of missing data.

We anticipated that the engagement would decline over time, consistent with other digital phenotyping studies and our previous report.^2^ Additionally, we expected that passive data collection would often correspond with active data collection from the same participants, but with higher collection rates ^7^.

### Computing data missingness and data completeness

We considered data missingness of accelerometer and GPS data at the day, hour, and minute levels. Consistent with the different “mechanisms of missingness,” we anticipated data missingness based on the design of our study. By design, accelerometer data are collected for 30 seconds and then not collected for 30 seconds.^18,22^ That is, if we hypothetically accounted for data at the second level, we can expect 50% data completeness with 50% data missingness for accelerometer data. Likewise, by design, GPS data are collected for 90 seconds and then not collected for 810 seconds. Therefore, at the hypothetical second level, we expect 10% data completeness with 90% data missingness for GPS data.

We used raw accelerometer and GPS data collected by Beiwe to examine missingness and completeness among all the participants at the three different timescales. For each data type, the total expected entries were calculated based on a 180-day study period, given that we considered “study completion” as 6 months (180 days) of data collection. The expected number of entries were computed for each of the three levels of granularity: day, hour, and minute.

Specifically, the expected entries should be as follows:

● Day level: 180 days
● Hour level: 180 × 24 hours = 4,320 hours
● Minute level: 4,320 × 60 minutes = 259,200 minutes

The raw data for each participant was downloaded and organized into individual folders based on their assigned Beiwe IDs. Within these folders, data were categorized by type, for example, with separate folders for accelerometer and GPS data. In each of these folders, the raw data was stored as CSV files, with each file name indicating the collection date and hour. A timestamp column in each file provided precise data collection times down to the millisecond.

For each timescale, we counted the actual number of unique entries. Consistent with previous studies^18,23^, if a single entry was observed at a specific timescale (a given day, hour, or minute), that time unit was counted as “collected.” The missingness (in %) for each participant at different levels was calculated using the formula:

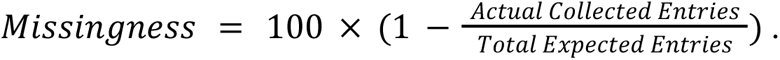

The completeness (in %), which is complementary to the missingness percentage, was calculated as:

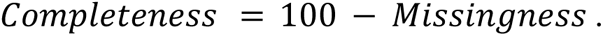

Here completeness represents the proportion of expected data that has been successfully collected. For participants whose data extended beyond 180 days, only the first 180 days were included in the missingness and completeness computations, and the rest were excluded.

### Trends and distributions in data completeness

To assess the distributions of data completeness across different timescales, we plotted histograms of the completeness percentages (see Figure 2). We combined the histograms for the day, hour, and minute levels into a single plot for each data type (accelerometer and GPS data) and used different colors to compare the timescales. To facilitate comparisons among these distributions, we also calculated and plotted the median completeness rates at each level for both data types, summarizing the central tendencies of data completeness across the timescales.

**Figure 2.**
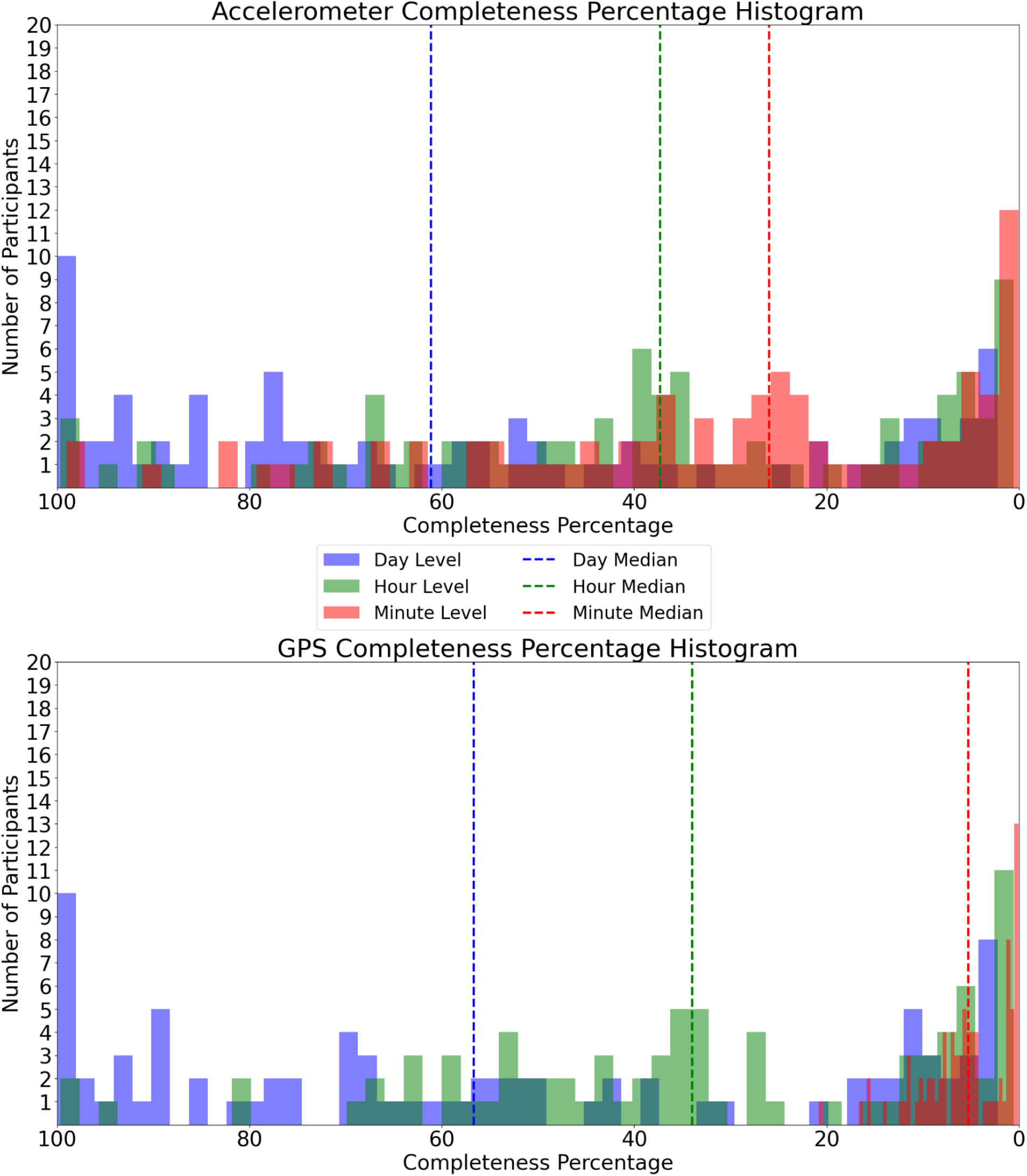
Impact of Timescale on Data Completeness (n = 85 participants). The histograms show the completeness percentages for accelerometer (top) and GPS (bottom) data across day, hour, and minute levels. As the timescale shortens, data completeness decreases, evidenced by the rightward shift of the distributions and the median lines.

### Impact of data missingness on summary statistics

We examined the relationship between hour-level and minute-level data missingness and day-level summary statistics generated by Forest. Specifically, we analyzed the impact of missing data on the day-level summary statistics for three measures: cadence (an accelerometer measure), home time, and number of significant locations (both GPS measures) (see Figure 3). Cadence was defined at the day level using the Forest Oak (forest.beiwe.org/en/latest/oak.html). Home time and number of significant locations were defined using Forest Jasmine (forest.beiwe.org/en/latest/jasmine.html). Home time is the span of time (in hours) spent at home between 8:00 PM and 8:00 AM each day,and home is defined as the most frequently visited significant location during these hours; significant locations are distinct pauses which are at least 15 minutes long and 50 meters apart.

**Figure 3.**
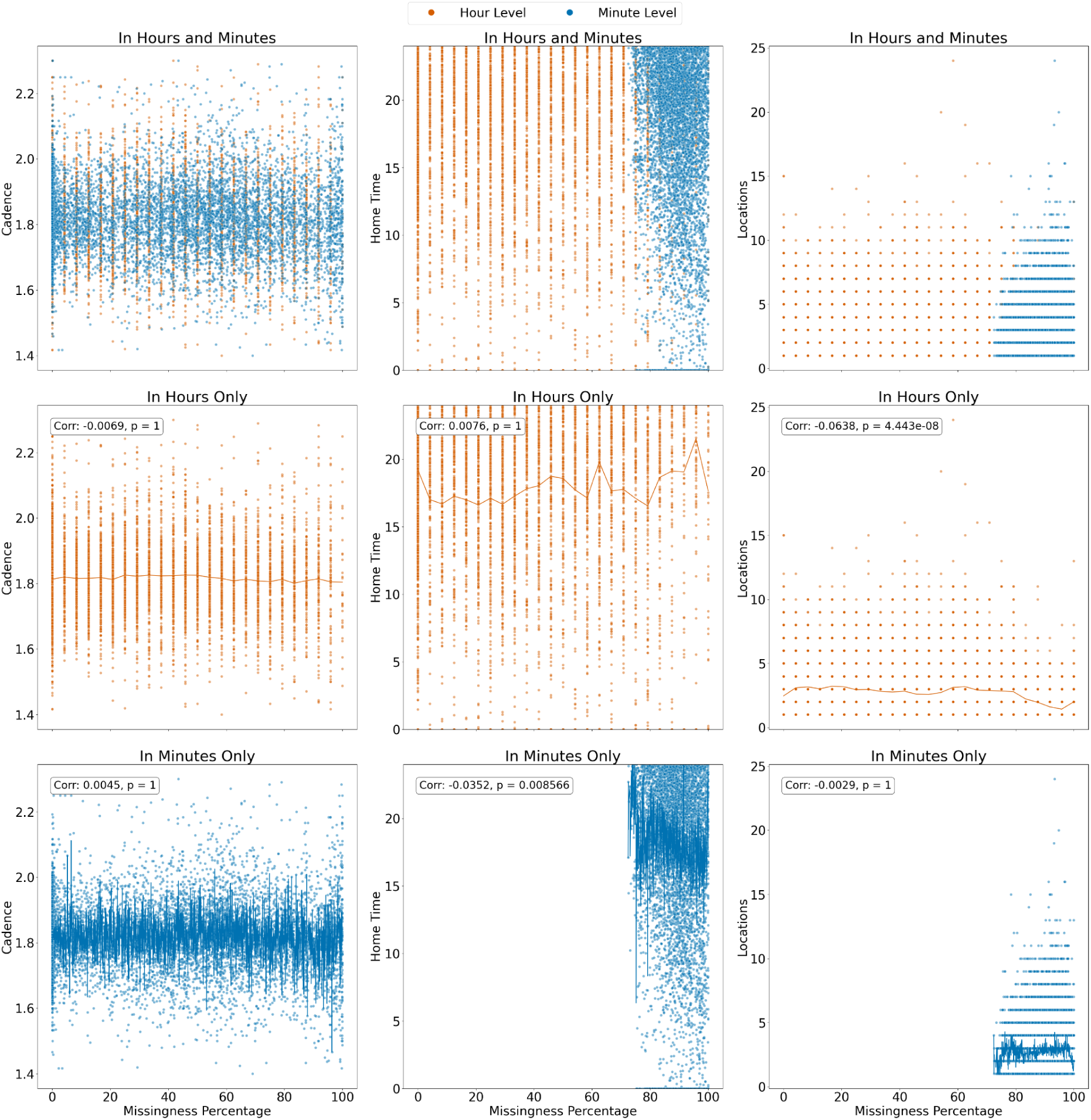
Association between data missingness and Forest measures at hourly and minute levels. These scatter plots show and compare the relationships between missingness percentages (x-axis) and daily summary statistics (y-axis) for the non-imputed accelerometer measure (cadence, left) and imputed GPS measures (number of hours spent at home, middle, and number of significant locations, right). Orange points represent hourly data, and blue points represent minute-level data. Very weak associations between missingness and both imputed and non-imputed measures are observed across all domains. Bonferroni correction for multiple comparisons was used to adjust displayed p-values, which were based on two-tailed tests. Significant relationships are identified with corrected p-values below 0.05.

It should be noted that while GPS data is imputed, accelerometer data is not, as no imputation methods exist for raw accelerometer data to our knowledge.^18^ This analysis allowed us to test whether the amount of missing data influences the summary scores provided by Forest. Ideally, there should be no relationship between missingness and summary measures intended to capture functional status. The GPS imputation method was designed to fill in missing data in a way that preserves the overall statistical properties of the raw data and does not introduce any unintended biases or relationships ^24,25^. Thus, if the imputation method is functioning correctly, we would expect no relationship between data missingness and the imputed GPS values.

To test the relationship between data missingness and the daily summary scores, we computed a Pearson correlation coefficient (*r*). This coefficient measures the strength and direction of a linear relationship between two continuous variables, ranging from -1 (perfect negative correlation) to +1 (perfect positive correlation), with 0 indicating no linear relationship. We also calculated the corresponding p-values to assess the statistical significance of these correlations. The p-value represents the probability that the observed correlation could have occurred by chance if there were no actual relationship (i.e., if the null hypothesis *r* = 0 is true). For this analysis, we used two-tailed p-values to account for the possibility of both positive and negative correlations. To account for multiple comparisons, we adjusted the p-values using the Bonferroni correction by multiplying each p-value by the total number of tests (6 in this case), as we conducted pairwise correlation computations for three variables of interest, with each variable being compared to the other two. A p-value below 0.05 after adjustment suggests that the correlation is unlikely to have occurred by chance, indicating a statistically significant relationship. We used Python’s ‘scipy.stats’ module to compute the Pearson correlation coefficients and the associated p-values.

### Analysis of variability in summary measures

To study the variability of summary measures throughout the 180-day study period, we computed the rolling standard deviation of two key measures: cadence and home time. The rolling standard deviation was calculated using a sliding window of 5 days, requiring all 5 days within the window to contain valid data points. Since there are 24 hours in a day, the values and the standard deviations for home time were logically capped at 24 hours. Hence, we applied a 24-hour threshold for home time as our criterion for identifying and excluding outliers. This method was applied to the data collected during the first 180 days of each participant’s study period. For participants who withdrew early, the analysis was conducted using the entire duration of their participation.

The resulting individual-level standard deviations were plotted over time for each participant, with color-coding based on the average percentage of missing data at the day level, where warmer colors (red) indicate higher missingness and cooler colors (blue) represent lower missingness (see Figure 4, left column). Gaps in the plots indicate periods where data sparsity – due to a high prevalence of missing values in the original data and, upon inspection, the removal of an outlier with a home time value of 115.76 hours on a certain date – resulted in insufficient valid data points for the standard deviation calculation. We also plotted the daily mean of the standard deviations throughout the study (black line) to assess their stabilization. More importantly, this analysis was designed to test whether the Forest GPS imputation quality improves over time, which could be indicated by a gradual trend of stabilization, as participants’ longer engagement in the study would allow the model to better impute their daily routines and patterns for the GPS data.

**Figure 4.**
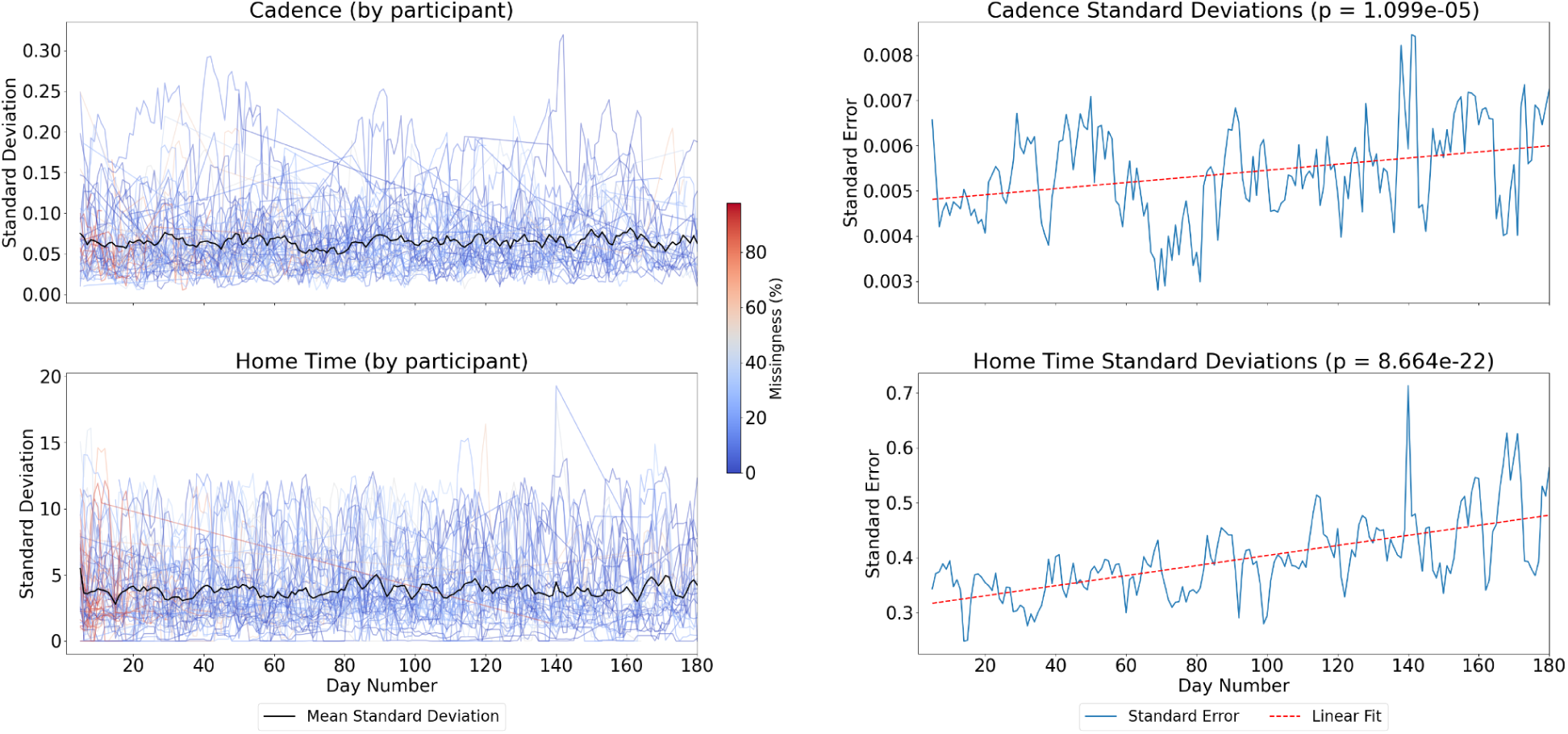
Standard deviation and standard error of summary statistics throughout the study period. This figure presents the individual-level rolling standard deviations (left column) and the population-level standard error of standard deviations (right column) for cadence (top row) and number of hours spent at home (bottom row) throughout the 180-day study period. The rolling standard deviations were calculated using a 5-day sliding window, requiring all 5 days to have valid data points. Colors in the left column represent the average day-level percentage of missingness, with black indicating the mean of the standard deviations per day. While individual variability is present in the standard deviations, the mean standard deviations suggest overall stabilization over time. Gaps in the lines on the left are due to data sparsity, where a large amount of missing values or outlier removal led to insufficient data for standard deviation calculation. In the right column, the standard error of the standard deviations reflects the variability of the standard deviations across the population of participants. A slight increasing trend is observed in the standard errors for both measures. The red dashed lines indicate linear fits, with p-values shown in the subplot titles. A p-value less than 0.05 suggests a statistically significant trend. Peaks in the standard error, particularly in the later stages of the study, may indicate periods of increased variability in participant data contribution.

In addition to these individual-level plots, population-level standard errors of standard deviations were computed and plotted (see Figure 4, right column). For each day, the standard error was calculated using the following formula:

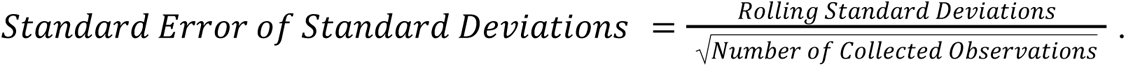

The standard error reflects the variability of the standard deviations across participants. Linear regression lines (red, dashed) were fitted to these standard error plots to examine any trends over time, with corresponding p-values indicating the statistical significance of the trends (significant if *p* < 0.05). These subplots aimed to assess whether the length of participants’ involvement in the study would influence their data contribution.

Moreover, the rolling mean count of valid data points for both the accelerometer and GPS measures was computed using a 5-day sliding window approach, the same method used for the standard deviation calculation (see Supplementary Figure 1).

### Impact of data missingness on regression model stability

We examined how varying levels of data missingness affect the stability and statistical significance of Ordinary Least Squares (OLS) regression models. Our analysis focused on the relationships between collected active data (PROMIS-29 survey subscores) and passive data (accelerometer-based cadence and GPS-based home time, analyzed separately). Minus the pain score (assessed as 0-10, 0 being no pain and 10 being the worst possible pain), the PROMIS-29 survey has 28 questions, each of which is categorized into seven subdomains: Anxiety (ANX), Depression (DEP), Fatigue (FAT), Physical Function (PF), Pain Interference (PI), Sleep Disturbance (SLP), and Ability to Participate in Social Roles and Activities (SOC). The selection of cadence and home time as variables of interest from the many available daily summary measures (see forest.beiwe.org for details) was made in consultation with the Beiwe team based on their robustness to data missingness. Cadence was chosen for its ability to be accurately computed with minimal data. For instance, an accurate measure of a participant’s cadence on a given day can be derived from just ∼2 minutes of data, compared to daily step count, which by definition represents an entire day’s activity. Likewise, since being at home is a relatively common event, home time is easier to impute over time than, for example, the number of significant locations visited.

Prior to model building, we first computed the hour-level missingness percentages for each participant for both accelerometer and GPS data, using the same formula as previously described.

Participants were filtered based on their hour-level missingness, with thresholds ranging from 0% to 100%. Two different approaches were taken: 1) each interval included data up to the corresponding missingness percentage (i.e., ≤1%, ≤2%, ≤3%, etc.) (see Figure 5 and Figure 6) and 2) each 20% missingness range was considered separately, not cumulatively (i.e., 0-20%, 20-40%, 40-60%, 60-80%, and 80-100%) (see Supplementary Figure 2 and Supplementary Figure 3). We used two strategies to test 1) whether cumulatively adding more data would change the OLS model results and 2) whether a specific missingness range would change the OLS model results.

**Figure 5.**
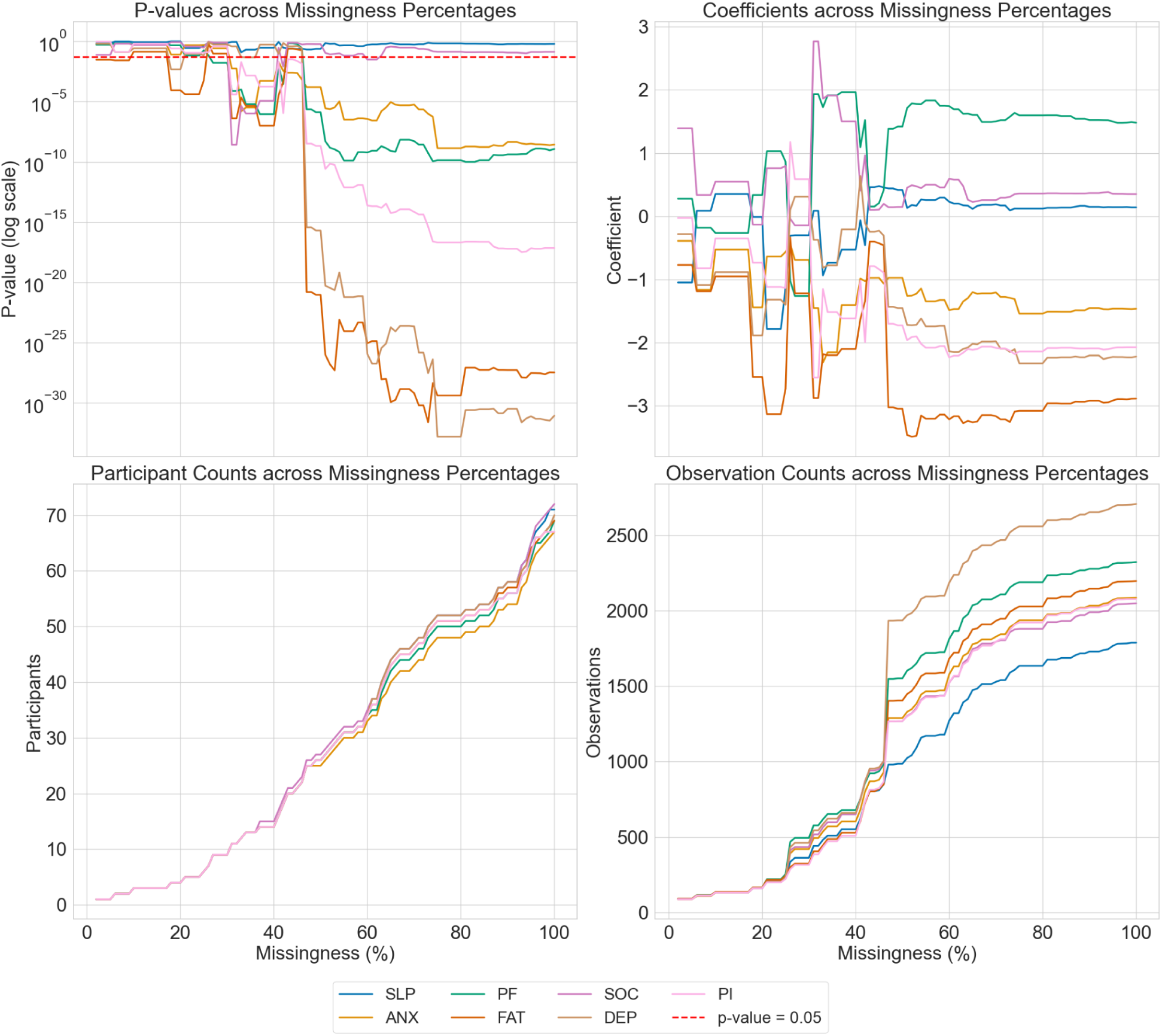
Cadence relationships with functional status across varying levels of hour-level data missingness. The Ordinary Least Squares (OLS) regression models for cadence (accelerometer data) show that the coefficients start to stabilize after 40% missingness, while the p-values (log scale) for significant PROMIS categories begin to decrease beyond this threshold. Significant relationships are identified with p-values below 0.05 (red dashed line). The analysis covers missingness intervals from 0% to 100%, with each interval including data up to the corresponding missingness percentage. P-values and coefficients are displayed in the top subplots, while participant and observation counts are shown in the bottom ones.

**Figure 6.**
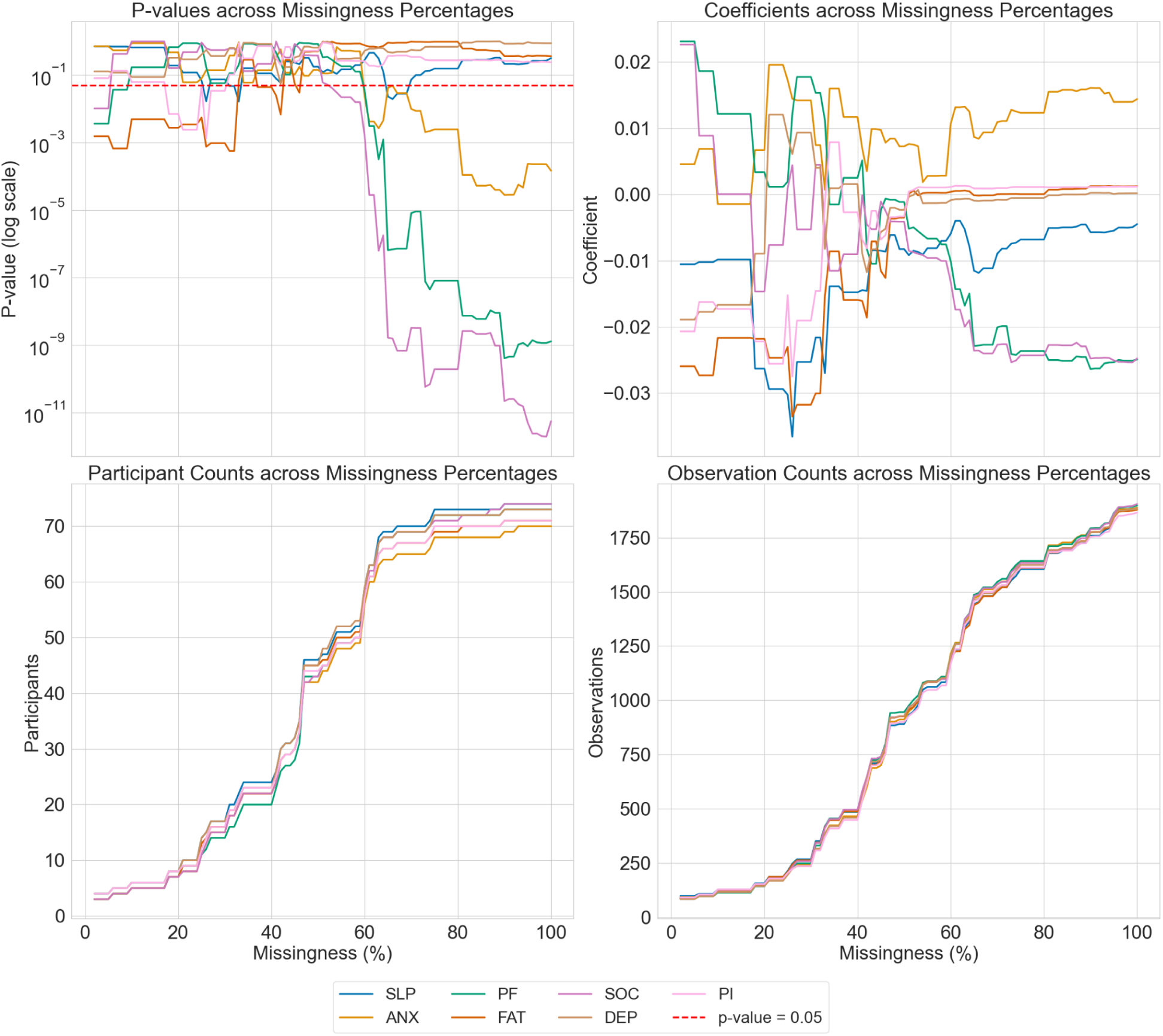
Home time relationships with functional status across varying levels of hour-level data missingness. The OLS regression models for home time (GPS data) show that while the coefficients begin to stabilize after ∼60% missingness, the p-values (log scale) for three of the PROMIS categories start to decrease and show significance. Significant relationships are identified with p-values below 0.05 (red dashed line). The analysis covers missingness intervals from 0% to 100%, with each interval including data up to the corresponding missingness percentage. P-values and coefficients are displayed in the top subplots, while participant and observation counts are shown in the bottom ones.

OLS regression models were then fit separately for each missingness interval and and for each PROMIS-29 category. The equation for the models is as follows:

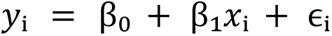

where *y*_i_ represents the PROMIS-29 subscore, *β*_0_ is the intercept, *β*_1_ is the coefficient for the independent variable (*x*_i_), which is either cadence or home time, and *ε*_i_ is the error term for observation *i*.

For each model, a constant term was included to represent the intercept. The models were assessed by examining the p-values and coefficients across the missingness percentages. Statistically significant relationships were identified where p-values fell below 0.05 (indicated by a red dashed line). For visualization purposes, the p-values were presented on a log scale. We also counted the number of participants and observations included in each model.

### Relationship between data missingness and functional status

We next aimed to identify potential data missing not at random (MNAR) patterns in the data set, which could suggest that functional status influences participants’ engagement with the study or the completeness of their data (see Figure 7).

**Figure 7.**
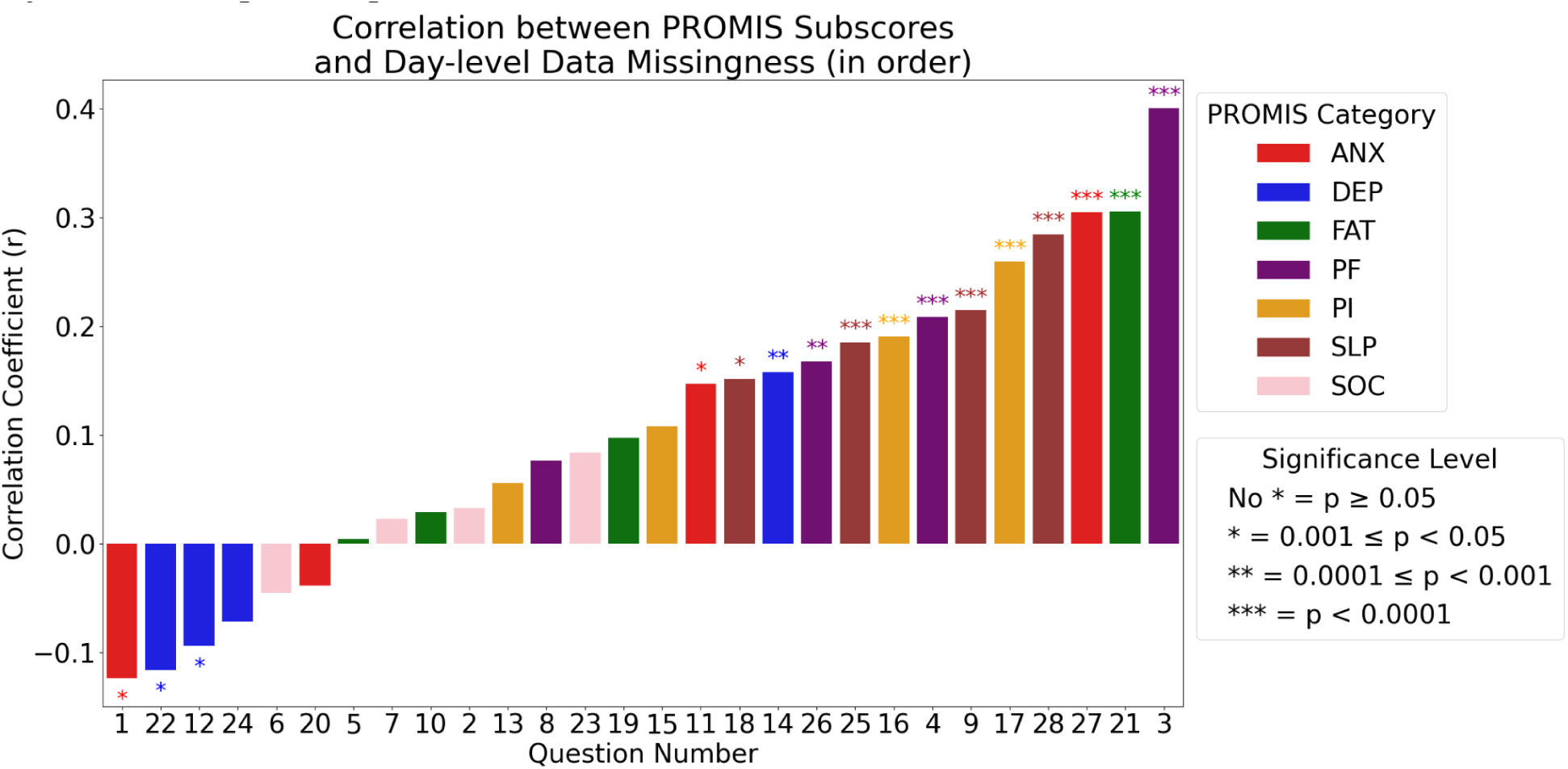
Is data missingness associated with functional status? Using the Pearson correlation coefficient (r), this bar plot examines the strength and direction of the relationship between participants’ functional status, measured with the PROMIS-29 subdomains, and their day-level PROMIS data missingness. The results, ordered from the most negative correlations on the left to the most positive on the right, suggest that functional status may be linked to data missingness, pointing to potential data missing not at random (MNAR) patterns. Two-sided p-values were computed to assess the significance of the correlations, and Bonferroni correction was applied to adjust for multiple comparisons.

To explore this, we calculated Pearson correlation coefficients (*r*) between standardized PROMIS subscores, which are on an ordinal scale from 1 to 5, and day-level PROMIS data missingness. The missingness for each participant was calculated using the formula described earlier. Although Pearson’s correlation is typically used for continuous data, we applied it here to evaluate the strength and direction of the linear relationship between these two variables, while acknowledging the ordinal nature of the scores. Specifically, we analyzed the correlation coefficients by PROMIS questions and categories.

We also computed the corresponding p-values using a two-sided test, accounting for the possibility of both positive and negative correlations. The null hypothesis tested by the p-values was that there is no linear relationship between the two variables. To address the issue of multiple comparisons, we adjusted these p-values using the Bonferroni correction, which involved multiplying the original p-values by the total number of tests. Given that there are 28 PROMIS questions, we conducted 28 separate correlation tests, resulting in a total of 28 comparisons.

### Visualizing Data Missingness and Uncertainty

Clinicians regularly use data visualizations to estimate means and trends across time series data; e.g. a series of blood pressure readings over many years is used to identify trends or changes in trends associated with the addition of an anti-hypertensive. Standard clinical data, however, are binary–either the value is present or it is not; “data missingness” as a concept does not exist because those data are simply not presented to the clinician (if an EKG has 60% missing data, the EKG is simply repeated). Data that are folded into clinical inference and decision come from machines and sensors that have calibrated thresholds for accuracy. The fact that digital phenotyping data can have a daily summary measure (e.g., home time) while representing varying amounts of missing data suggests that these dependencies should be readily apparent in any data visualization. In other words, visually communicating the extent of data missingness represents a responsibility of the data team as a larger amount of data missingness effectively communicates an added layer of uncertainty in measurement. Methods to visualize data missingness are needed if digital phenotypes are to guide clinical inference and decision.

While not directly aimed at clinical work, a compelling series of analyses by Sarma et al. ^26^ systematically analyzed how the (human) accuracy varied as a result of different data visualization methods in two tasks: average estimation and trend estimation. The goal of each method was to communicate to the viewer varying levels of uncertainty related to missing data (data missing at random). Sarma et al’s report an “almost deterministic” relationship of showing imputed estimates across six types of visualization methods, which included baseline (no imputation shown), mean point estimate, confidence interval (95%), probability density plots, gradient (of 95% CI), and hypothetical outcome plots (HOPs) of predicted imputed values.

When estimating averages, uncertainty representations may reduce bias but at the cost of decreasing precision.When estimating trend, only hypothetical outcome plots may lead to a small probability of reducing bias while increasing precision whereas CI, density, and gradient did not consistently reduce bias nor improve precision. Critically, at higher levels of data missingness, HOPs consistently improved precision as well as reduced bias. Overall, Sarma et al report that a participants ability to estimate mean and trend depends on the complexity of the visual data they need to digest: the more marks a plot has, the harder a viewer must work to consolidate and encode uncertainty information, thus decreasing precision and increasing bias. In other words: simpler is better, but not too simple.

Based on this “simpler is better” principle, we demonstrate uncertainty information by color coding data based on the extent of missing data. For an individual participant, we illustrate daily values with <25%, 25-50%, 50-75%, and ≥75% missing data (at the hour level) (see Figure 8).

**Figure 8.**
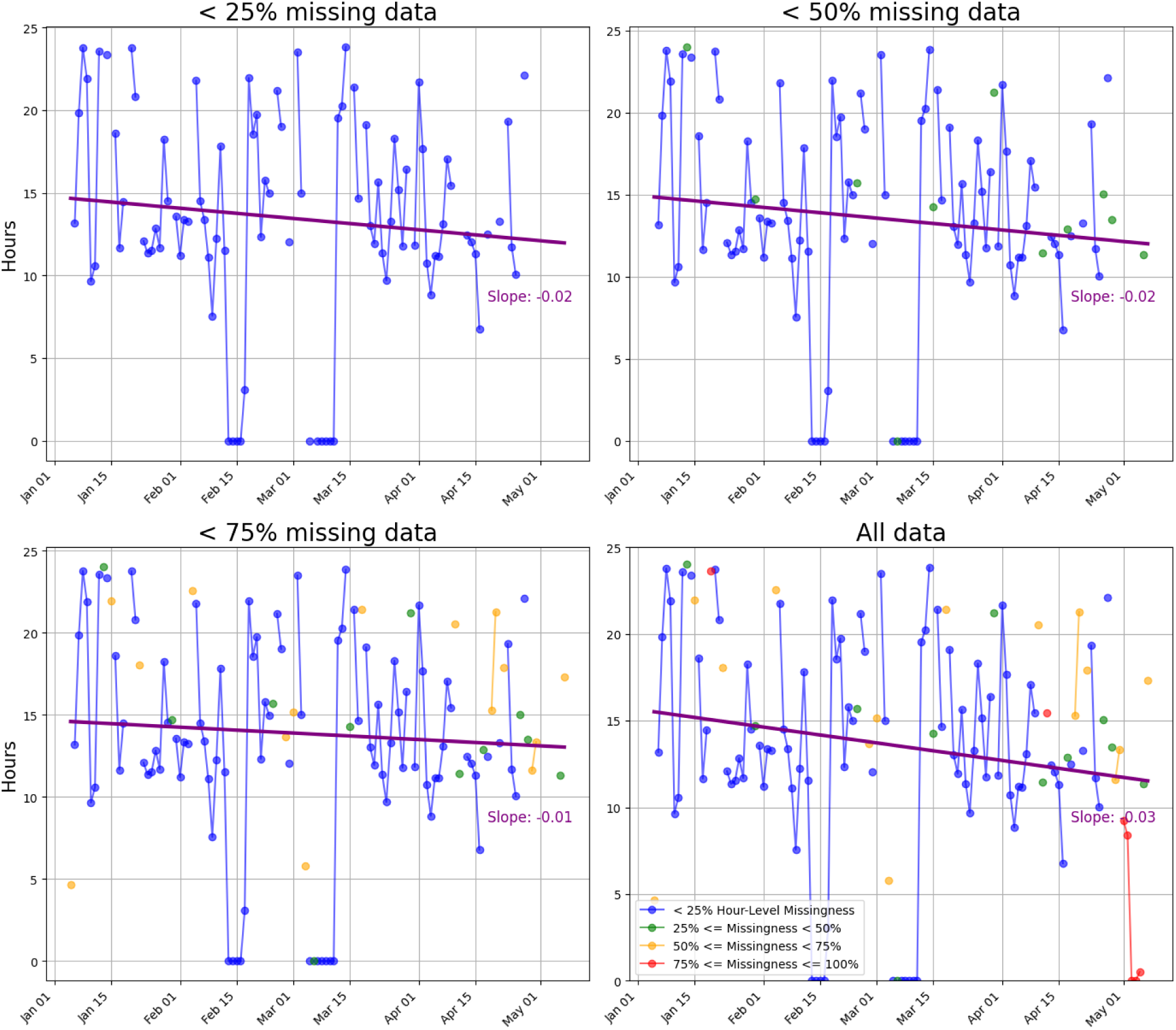
Individual patient visualization of hour-level data missingness. Because daily summary scores might have informative signals, even in the presence of missing data, we determined to not simply discard missing data based on a threshold. Instead, to communicate the uncertainty underlying a daily summary score (shown along the y-axis), we color coded brackets of data missingness (blue, 0-25%; green, 25-50%; yellow, 50-75%; red, ≥75%) and progressively added data brackets to each quadrant (left-right, top-bottom). Regression line noted with the solid purple line, without a notable difference in slope (labeled in purple text) across the quadrants suggesting that for this patient, there was minimal difference in the mean or the trend of home time related to data missingness.

## Results

### Relationship of data completeness with study duration

Figure 1 illustrates the data completeness for passive data (accelerometer and GPS measures) and active data (PROMIS-29 survey responses) over the 6-month study period, from day 1 to day 180 (or from hour 1 to hour 4320), at both the day and hour levels. Since the PROMIS survey questions are prompted to participants daily, hour-level PROMIS data missingness was considered as missing by design and was not included in our analyses.

The day-level completeness for the accelerometer data (Figure 1, top panel, green line) starts high but declines within the first month (the first 30 days or the first 720 hours) due to several participants withdrawing early (marked by yellow dots). Completeness then stabilizes around 80% for most of the study period, with an increase towards the end. We attribute this observed increase in data completeness to an artifact of fewer participants remaining in the study, inflating the percentages (e.g., a single participant providing data would constitute 100% of participants). Similarly, the GPS data (Figure 1, middle panel, green line) follows a similar pattern, with high initial completeness, a decline in the first month, stabilization around 80%, and a late upward trend.

In contrast, the hour-level completeness for the accelerometer data (Figure 1, top panel, blue line) shows more variability. The initial high completeness quickly drops, indicating early withdrawals (marked by red dots), followed by fluctuations throughout the 4320-hour study, with a gradual improvement towards the study’s end. The GPS data (Figure 1, middle panel, blue line) mirrors these patterns. Overall, day-level completeness is higher than hour-level completeness for both the accelerometer and GPS data.

For PROMIS data (Figure 1, bottom panel, green line), day-level completeness is universally lower than both the accelerometer and GPS data, stabilizing between 40% and 50%, with a substantial, artifactual increase near the end of the study. Despite this, the day-level patterns are consistent across the accelerometer, GPS, and PROMIS data. The relationship between active and passive data collection can be understood in two ways: a) active data requires participants to take the initiative to input their responses, while passive data could be automatically uploaded to the Beiwe platform, and b) when passive data is missing, active data is likely missing as well.

### Data completeness differences by timescales

The patterns of data completeness for both accelerometer and GPS data were analyzed across day, hour, and minute levels. Overall, as the timescale decreased, the computed data completeness also decreased. We discuss the specific trends observed in accelerometer and GPS data separately below.

For both accelerometer and GPS data, more participants exhibited lower completeness percentages at the hour and minute levels. At the day level, by contrast, the number of participants with data collected initially decreased but then increased toward the lower end of the completeness spectrum. Additionally, more participants showed 100% data completeness than near-zero completeness at the day level.

In the accelerometer data (Figure 2, top panel), 10 participants achieved 100% data completeness at the day level, indicating no missing data throughout the 6-month period. However, no participants had 100% completeness at the hour or minute levels, though 3 participants at the hour level and 2 participants at the minute level had completeness very close to 100%.

For the GPS data (Figure 2, bottom panel), 10 participants also achieved 100% completeness at the day level, and 2 participants showed near 100% completeness at the hour level. However, the maximum completeness at the minute level was approximately 20%, with nearly all participants having less than 20% completeness.

Median completeness values further illustrated the decline in completeness with decreasing timescale. For accelerometer data, the median completeness was about 60% at the day level, 37.29% at the hour level, and 25.94% at the minute level. GPS data showed lower median values across all levels compared to accelerometer data, with 56.67% at the day level, 33.96% at the hour level, and 5.27% at the minute level.

These findings underscore the challenge of maintaining data completeness in high-frequency data collection, where finer granularity (i.e., minute level) is associated with lower completeness compared to broader granularity (i.e., day level). These findings further underscore the need to articulate at which timescale data completeness and missingness are being accounted for.

### Association between missingness and Forest measures

At the hour and minute levels, we were able to define a relationship between data missingness and the measures of accelerometer-based cadence and GPS-based home time and number of significant locations visited.

More specifically, for the accelerometer data (Figure 3, left column), the analysis demonstrates no discernible association between missingness and cadence at either the hourly or minute levels, with the data points scattered uniformly across the range of missingness percentages. The hour-level correlation coefficient (*r* = -0.0069, *p* = 1) indicates a very weak and statistically non-significant negative association. Similarly, the minute-level data reveals an extremely weak and non-significant positive association (*r* = 0.0045, *p* = 1). These results suggest that data missingness does not significantly impact the daily accelerometer-based cadence.

For the GPS data (Figures 3, middle and right columns), the plots reveal a more distinct pattern. At the minute level, data clusters at higher missingness percentages. The hour-level data, however, remains relatively evenly spread. For home time, the hour-level correlation (*r* = 0.0076, *p* = 1) suggests a very weak and non-significant positive association, while the minute-level data shows a statistically significant though weak negative association (*r* = -0.0352, *p* = 0.008566), indicating that higher missingness corresponds to shorter home time. Regarding number of significant locations, the hour-level correlation coefficient (*r* = -0.0638, *p* = 4.443e-08) also indicates a statistically significant weak negative association, while the minute-level data shows a near-zero and non-significant negative association (*r* = -0.0029, *p* = 1). This suggests that higher missingness has had little impact on the number of significant locations visited. Given these observations, the overall associations are weak, indicating minimal influence of missingness on the Beiwe-imputed GPS measures.

In general, the results indicate that data missingness, whether at the hour or minute level, does not significantly affect either imputed or non-imputed measures. This suggests that, regardless of imputation, data missingness has minimal impact on these measures, reinforcing the robustness of both imputed and non-imputed summary statistics against varying levels of missing data.

### Impact of missingness on variability in summary measures

Figure 4 presents the individual-level rolling standard deviations (left column) and population-level standard errors of standard deviations (right column) for cadence (top row) and home time (bottom row) over the 180-day study period. The individual-level plots illustrate the relationship between the standard deviation of daily summaries (y-axis), data missingness (line color, except for the black line, which represents the mean standard deviation), and study duration (x-axis). The population-level plots reflect the variability of these standard deviations across participants, with the red dashed line representing the linear fit to the standard errors and p-values displayed in the subplot titles.

The individual-level results indicate that while the standard deviations of cadence and home time remain relatively stable over time, persistent fluctuations are observed among individuals throughout the study. In both plots, participants with higher data missingness tend to appear more frequently during the first 60 days of the study. This could be explained by the study design, where participants were dismissed after more than two consecutive months of low completion. Variability in the standard deviations among individuals is more persistent throughout the study period, regardless of the level of missingness, and is notably higher for home time compared to cadence. This variability in home time is reflected by the more pronounced fluctuations in its mean standard deviation, as indicated by the black line. Gaps in the plotted lines emphasize the impact of missing data on the stability of the summary measures, particularly in the early stages of the study when data collection may have been less consistent.

Overall, the visualization suggests that participants would engage more consistently over time, yet the variability in their summary measures remains about the same, particularly for cadence. However, variability in home time remains more noticeable, likely due to the nature of the data and the extent of missingness in the raw data. Contrary to expectations, no trend of decreasing mean standard deviation is observed over time, indicating that the imputation method has not really improved as the study progresses. Additionally, there is no clear relationship between higher levels of data missingness (represented by the blue-red color bar) and greater variability in either measure. For instance, a participant with lower data missingness could show a larger standard deviation in the imputed home time measure than another participant with more missing data, as evidenced by the presence of highly positioned blue lines compared to lower red lines in both plots.

Supplementary Figure 1 further supports this finding by showing the rolling mean counts of valid data points for accelerometer and GPS measures across participants throughout the study period. The mean counts remain relatively stable and high over time, indicating consistent data contribution from participants at the population level, despite individual differences in missingness. For more details, please refer to the supplementary figure.

The population-level results provide a broader perspective on the variability of the standard deviations across participants. For both cadence and home time, a small increasing trend in the standard errors can be observed, as evidenced by the positive slopes of the red dashed linear regression lines. Additionally, both p-values being well below the 0.05 threshold confirm that these positive relationships are statistically significant, with the p-value for home time being much smaller than that for cadence. This suggests that variability in the standard deviations tends to increase slightly as the study progresses. Peaks in the standard errors are particularly evident toward the end of the 180-day study period, especially in home time. These peaks may reflect periods of increased variability in participant data contribution and/or a decreased number of observations collected per day, given the formula used to calculate the standard errors.

Consistent with the previous figure, we can conclude that, regardless of whether the measures were imputed (GPS data) or not (accelerometer data), the findings are similar between them. This shared pattern suggests that both imputed and non-imputed measures exhibit comparable trends in variability over time.

### Impact of hour-level data missingness on regression model stability

A total of 81 participants had available hour-level accelerometer missingness percentages, with a mean missingness of 63.46%, a median of 62.75%, and a standard deviation of 26.81%, indicating considerable variability across participants. Notably, 66.67% of participants had more than 50% missing accelerometer data at the hourly level. Similarly, 82 participants had available hour-level GPS missingness percentages, with a mean missingness of 68.23%, a median of 65.94%, and a standard deviation of 23.88%. A slightly higher proportion of participants (70.73%) had more than 50% missing GPS data.

For both accelerometer-based cadence and GPS-based home time, the coefficients (Figures 5 & 6, top right panels) of the OLS models fluctuate substantially at lower levels of missingness. The coefficients for cadence begin to stabilize after 40% missingness, while those for home time show stabilization after approximately 60% missingness. This pattern suggests that as hour-level missingness increases beyond these thresholds, the models’ predictions for these variables become more consistent and reliable.

For cadence, the p-values (Figure 5, top left panel) for several PROMIS categories, including PF, PI, ANX, FAT, and DEP, decrease and remain statistically significant beyond the 40% threshold, except for SLP and SOC, which are insignificant at higher levels of missingness. Notably, the p-values for SLP remain above 0.05 throughout the missingness spectrum, illustrating its lack of statistical significance. In contrast, for home time (Figure 6, top left panel), after the 60% threshold, only ANX, PF, and SOC become statistically significant, while the other PROMIS-29 categories have p-values above 0.05, indicating that they are not statistically significant.

Furthermore, the number of participants included in the analysis (Figures 5 & 6, bottom left panels) increases with the percentage of hour-level missingness. This trend suggests that as more missing data is allowed, more participants can be included in the analysis. However, it is important to note that the quality and informativeness of the data may decrease with higher levels of missingness. The number of observations (Figures 5 & 6, bottom right panels) also increases. Notably, while the number of observations across missingness percentages diverges for cadence after the 40% threshold, the number of observations for home time tends to follow a consistent pattern across all PROMIS-29 categories. Although the inclusion of additional observations may seem beneficial, it emphasizes the necessity of carefully addressing missing data to maintain the robustness and accuracy of our findings. This also suggests that higher levels of missingness at specific timescales may reduce the insightfulness of our results, as different PROMIS-29 categories converge and lead to similar modeling outcomes when missingness percentages are high.

Supplementary Figures 2 & 3 further elaborate on these trends. Noticeably, though the modeling results differ between cadence and home time, the 40-60% missingness range emerges as a critical turning point for both variables, reinforcing the need to consider the impact of missingness when interpreting these outcomes. For more details, please refer to the supplementary figures.

### Relationship between day-level data missingness and functional status

We examined the relationship between participants’ functional status and day-level PROMIS data missingness. There were altogether 82 participants whose day-level PROMIS data missingness was computed, with a mean of 70.81%. The median was 75.56%, the standard deviation was 24.88%, and 76.83% of the participants had more than 50% data missing in their daily PROMIS responses. The results revealed varying degrees of correlation across different PROMIS questions and categories, suggesting that functional status may influence the likelihood of missing data.

Data missingness in daily surveys can occur under contrasting conditions of physical, mental, or social health. Both improvements and declines in health can reduce participants’ engagement with the surveys: those who feel better may become more active and thus neglect the surveys, while those experiencing worsening health may struggle with the physical or emotional burden of participation. This dual scenario highlights the complex relationship between health status and survey engagement, where both positive and negative changes in functional status can lead to increased missing data for different reasons.

Consider the highest and second-highest correlation coefficients shown in Figure 7. The strongest correlation is observed in response to Question 3 (PF), which asks, “Are you able to go for a walk of at least 15 minutes?” with responses ranging from 5 (“Without any difficulty”) to 1 (“Unable to do”). The positive correlation (*r* = 0.40, *p* = 7.13e-37) indicates that as day-level data missingness increases, the corresponding score tends to be higher, and this correlation is highly statistically significant. This suggests that participants experiencing improvements in physical function (e.g., when they are able to walk more easily), may become less inclined to complete daily surveys. They may be too occupied with daily activities like walking or socializing and thus forget to answer the question, or they may perceive less need to engage with the surveys, leading to higher levels of data missingness.

On the opposite, the second-highest correlation is given by Question 21 (FAT), which asks, “During the past 7 days… How run-down did you feel on average?” with responses ranging from 5 (“Very much”) to 1 (“Not at all”). Similarly, this significant positive correlation (*r* = 0.31, *p* = 1.65e-14) means that as day-level data missingness increases, so does the reported score.

Participants who feel more tired are also more likely to miss completing their daily surveys. Fatigue can diminish motivation and initiative, leading participants to prioritize rest or other coping mechanisms over maintaining consistent survey participation, thereby increasing data missingness.

A comparison between the third-highest and the lowest correlation coefficients in Figure 7 further illustrates the complexity of the relationship. The third-highest correlation (*r* = 0.30, *p* = 2.09e-17) comes from Question 27 (ANX), which asks, “In the past 7 days… I felt uneasy.” with responses ranging from 5 (“Always”) to 1 (“Never”). The lowest correlation (*r* = -0.12, *p* = 1.48e-02) is observed with Question 1 (ANX), which asks, “In the past 7 days… I felt fearful.” with the same response options. Both correlations are statistically significant but could be interpreted from two completely different perspectives, given that one is positive and the other is negative. For Question 27, a higher score indicates greater anxiety, which may distract participants and lead them to prioritize addressing their worries and unhappiness. As a result, the participants may forget to answer the daily surveys, leading to more missing data. Conversely, for Question 1, a lower score suggests improved mental health, which may lead participants to deprioritize survey completion, also resulting in higher levels of missing data.

Additionally, the p-values indicate that the top thirteen correlations are statistically significant, with the significance decreasing as the correlation coefficients decrease. The three most negative correlations all have p-values between 0.001 and 0.05, underscoring their statistical significance.

Overall, these results emphasize the complex interplay between functional status and data collection. Both positive and negative changes across different dimensions of health can contribute to increased daily survey data missingness. This suggests that participants’ functional status may influence their engagement with the study, potentially leading to data that is missing not at random (MNAR) in a clinically significant way.

### Visualizing Missing Data at Patient-Level

Consistent with our larger goal of presenting digital phenotyping data in a way that might guide clinical inference and clinical decision, we researched and adapted a method for data visualization that allows a hypothetical clinician to view a patients’ data in the context of increasing levels of data missingness, shown in Figure 8 with increasing amounts of data, depending on data missingness level. We present this illustration simply as a possibility based on the “simpler is better” principle described by Sarma et al.^26^

## Discussion

Missing data are a reality of digital phenotyping studies of clinical populations, especially over a protracted timeframe. For digital measures of functional status to enter clinical practice, data missingness must be accounted for and managed. We take a microscope to our real-world, longitudinal digital phenotyping data to understand data missingness in concrete, clinical terms. We show how accounting for missing data at the day, hour, and minute levels can lead to quite different distributions of data missingness over time. Notwithstanding these differences, Beiwe’s Forest toolset (Oak for accelerometer; Jasmine for GPS) imputed daily summary measures that were robust to variable levels of data missingness at the participant-level. At the group-level, we also show that linear statistical models are not robust to changes in data missingness, however these models show stability near the threshold of 40% (for accelerometer) to 60% (for GPS) data missingness. We provide evidence that functional status is related to and might impact data missingness. Therefore, we have shown that the timescale at which one accounts for missing data and the related threshold one uses to include a participant in a group-level design can impact the statistical results of the study. We now consider the larger goal of digital phenotyping and propose ways to manage missing data with an eye towards the clinical goals of the study.

Digital phenotyping promises to help us measure clinically relevant indices of patient function over time, including changes in function, importantly in real-time and in real world settings. As with any clinical instrument, the stability and boundary conditions of digital devices relate directly to inferences made from the resulting data. There is a large and growing body of research that compares the reliability of various consumer-grade digital devices. A recent review of 158 publications examining nine commercial wearable device brands (e.g., Fitbit, Apple Watch, Samsung, etc) showed that devices were accurate for measuring steps and heart rate in laboratory-based settings, concluding that more research was needed in various settings.^27,28^ Phenotypes derived from smartphone-based applications (as deployed in our study here) are known to vary as a function of how the application obtains its data (via native or web-based application^29^), and by smartphone sensors, platform, hardware, and OS.^30^ In our longitudinal study focused on detecting functional changes over time, we showed that measures and subsequent clinical inferences can further vary as a function of missing data. While missing data is a much-discussed topic in behavioral and social sciences, the traditional “missingness mechanisms” do not neatly apply to digital phenotypes.

The standard treatment of data missingness focuses on how to statistically overcome the missingness by imputing those missing values, with the appropriate statistical method depending on the “missing mechanism” that governs the missingness.^31^ In an excellent overview of the topic, Ibrahim et al. described the four most common imputation methods: Maximum Likelihood (ML), Multiple Imputation (MI), Fully Bayesian (FB), and Weighted Estimating Equations (WEEs).^31^ ML^32^ is widely used for data that is missing at random (MAR) and can theoretically be extended to handle missing not at random (MNAR) with additional modeling, though this is challenging. MI^16,33^, which effectively accounts for the uncertainty inherent in the imputation process, is also primarily designed for MAR but can be adapted for MNAR under certain conditions. FB methods are more flexible^31,32^ and can handle both MAR and MNAR scenarios, despite the difficulty of correctly specifying the model for MNAR. Lastly, WEEs^34^ are well-suited for data missing completely at random (MCAR) and MAR, only with some potential to handle MNAR, although this is often not reliable. In the clinical setting, one can generally assume that missingness falls under MAR or MNAR^35^, but MNAR could not only be the most challenging to handle, but also more prevalent, where the missingness is related to unobserved data itself. Although the aforementioned imputation methods might be adapted to address MNAR, doing so could be highly complex and not straightforward, often requiring intricate modeling that may not capture the full clinical picture.

The Beiwe team took a different approach by employing sparse online Gaussian Process (SOGP) for bidirectional imputation on a 3-dimensional sphere, incorporating several condition checks to enhance accuracy and computational efficiency.^24^ While our results show that the choice of SOGP is robust to variable levels of missing data, our analysis also suggests that this imputation method may smooth out some GPS signals, potentially leading to a loss of variability within individuals and across days; signal that has potential clinical value. For instance, SOGP’s direction adjustment function bends the flight – defined as a segment of linear movement – toward the destination with the highest probability, thereby almost eliminating the possibility of the trajectory starting in the opposite direction.^24^ In terms of smoothing a signal over time, our results validate that Beiwe’s SOGP does indeed stabilize GPS summary scores over a range of data missingness however perhaps at the expense of our specific goal of detecting inter-individual day-level changes in function.

Functional status and, in particular, pain can fluctuate moment-by-moment and day-by-day.^2,7^ The promise of digital phenotyping lies in its ability to track moment-by-moment changes and day-by-day trajectories, offering the potential for real-time interventions that can detect and mitigate functional decline. While valuable for managing missing data, highly effective imputation methods may inadvertently obscure patient-level variability, potentially leading to a loss of clinically meaningful information. That is, if we treat missing data purely as a statistical challenge rather than as a feature with clinical significance, we risk overlooking important signals that could portend a patient’s functional decline. This is particularly important regarding behavioral features of many disorders. Therefore, it is essential to account for missing data at multiple timescales (we recommend day, hour, and minute levels) and to consider missing data not only as a statistical challenge but also as a potential indicator of underlying clinical changes.

In the clinical world, unfortunately, no universally applicable methods of handling missing values can be recommended.^36^ Because of this, it is crucial not to bypass missing data and make clinical decisions in isolation. Instead, the conversation should integrate the clinical and statistical domains with an ideal approach carefully tailored to the specific clinical context and its potential implications for patient care. Furthermore, clinical decisions are often most guided by outliers, relatively infrequent and unexpected events in data. Detecting a *change* in function would, by definition, be more complex if data were imputed using any of the above tools. Therefore, while an imputation method may be statistically valid and facilitate subsequent group-level statistics, it may present a problem relevant to clinical decision: missing data itself could very well represent a signal or trajectory that is clinically relevant.

To address this issue, we propose that the existing framework of “missingness mechanisms” should be adapted to suit the specific needs of clinical digital phenotyping studies, described in Table 2. Our framework includes two previously proposed categories wherein data can be missing by design (MBD) and due to sensor non-collection.^18^ Based on the above work, we provide a rationale for further dividing “sensor non-collection” into three additional categories: technology failure, clinical change, and extraneousness. This framework is fit-for-purpose because it allows individual clinical studies to adapt what is considered “clinically relevant” and “extraneous” depending on their specific goal of data collection. Based on this framework, we emphasize the importance of accounting for missing data, as proper assessment is necessary to identify whether data are missing due to “clinical change” or “technology failure.” Longitudinal clinical deployment requires that “clinical change” and “technology failure” are events that must be mitigated, not simply imputed.

**Table 2.**
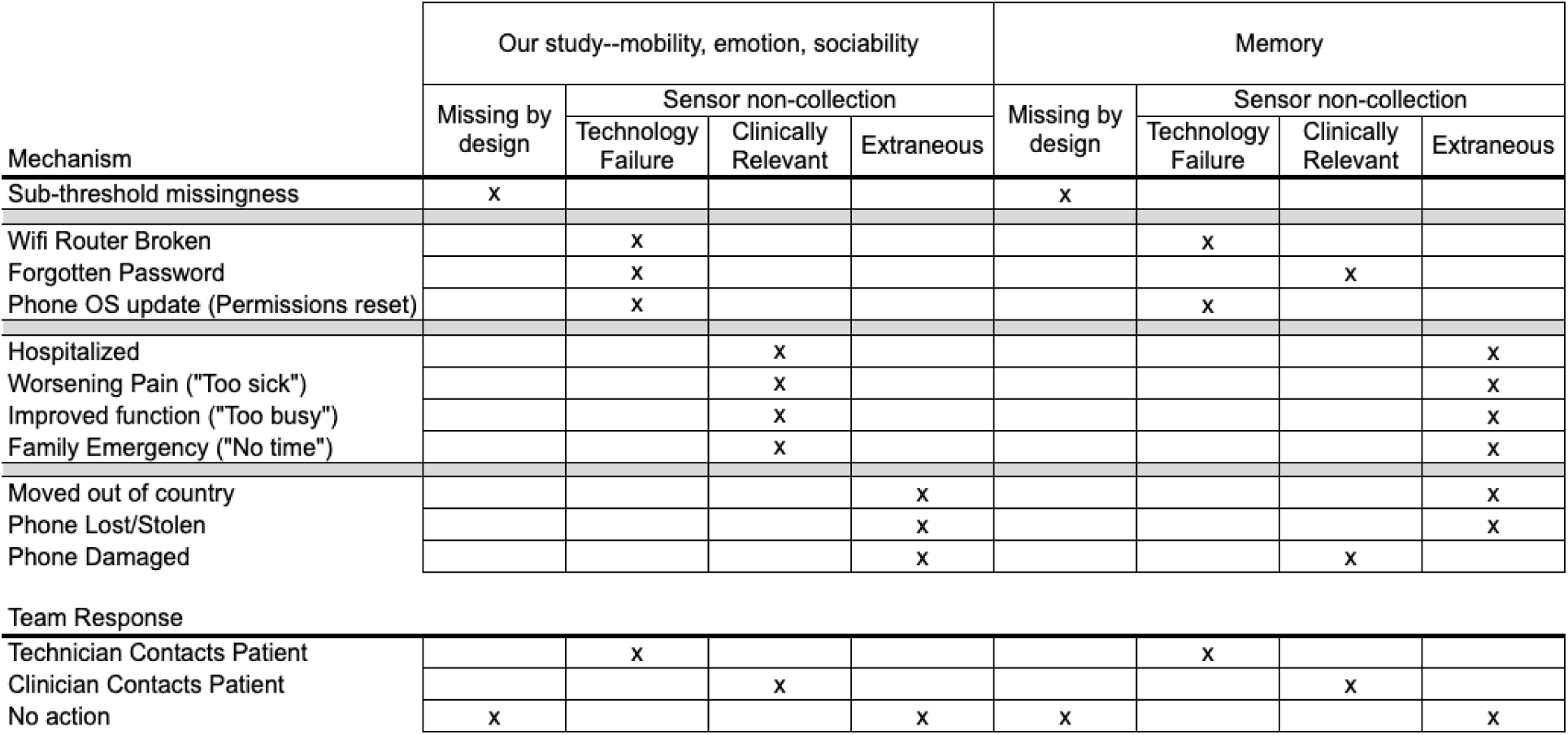
Clinical context is important when considering the “missingness mechanism.” We propose that mechanisms of data missingness include “Missing by design,” technology failure, clinically relevant, and extraneous. What is considered technology failure, clinically relevant, and extraneous depends on the clinical context within which the data are collected. We illustrate two studies: our own (focused on functional status), and a hypothetical study of memory. While a forgotten password or lost phone might, respectively, constitute a technology failure and extraneous reason for missing data in our study, forgetting a password and misplacing a phone might be clinically relevant events in a study of memory.

| Mechanism | Our study--mobility, emotion, sociability |  |  |  | Memory |  |  |  |
| --- | --- | --- | --- | --- | --- | --- | --- | --- |
|  | Missing by design | Sensor non-collection |  |  | Missing by design | Sensor non-collection |  |  |
|  |  | Technology Failure | Clinically Relevant | Extraneous |  | Technology Failure | Clinically Relevant | Extraneous |
| Sub-threshold missingness | x |  |  |  | x |  |  |  |
| Wifi Router Broken |  | x |  |  |  | x |  |  |
| Forgotten Password |  | x |  |  |  |  | x |  |
| Phone OS update (Permissions reset) |  | x |  |  |  | x |  |  |
| Hospitalized |  |  | x |  |  |  |  | x |
| Worsening Pain ("Too sick") |  |  | x |  |  |  |  | x |
| Improved function ("Too busy") |  |  | x |  |  |  |  | x |
| Family Emergency ("No time") |  |  | x |  |  |  |  | x |
| Moved out of country |  |  |  | x |  |  |  | x |
| Phone Lost/Stolen |  |  |  | x |  |  |  | x |
| Phone Damaged |  |  |  | x |  |  | x |  |
| **Team Response** |  |  |  |  |  |  |  |  |
| Technician Contacts Patient |  | x |  |  |  | x |  |  |
| Clinician Contacts Patient |  |  | x |  |  |  | x |  |
| No action | x |  |  | x | x |  |  | x |

Although imputed measures appear to be robust to data missingness, we provide evidence (see Figure 7) that the amount of data missingness and the reason for data missingness depends on the clinical context. In Table 2, we contrast our study of functional status with a theoretical study of memory. Consider that Beiwe requires patients to enter their password in order to input new data: if patients do not enter their password, no data can be collected. In our study, we observed that some patients not engaging in the active data collection had simply forgotten their password—an event that might be considered extraneous to our tracking of functional status. In a study of memory, however, participants forgetting passwords could be clinically relevant. Consequently, the approach each study would take to tackle this issue might differ: participants will either be contacted by a technician (in our study) or by a clinician (in the memory study). Therefore, the problem of missing data may require clinical expertise on the underlying reasons for the missing data.^37^ Especially when nowadays there is increasing interest in utilizing data science tools to help predict, diagnose, and prevent diseases, statisticians and clinicians should make decisions together about which assumptions are appropriate for the data at hand.^37^

Despite our proposal of a modified missingness framework, our study has several limitations that should be acknowledged. First, our relatively small sample size (n = 85 participants) limits the generalizability of our findings. We hope to overcome this limitation in future work with greater funding that permits a larger sample and welcome collaborations. We further note that, along the spectrum of population diversity, our sample skews largely white and predominantly women, which does not fully represent the broader population. Research has shown that digital device usage for health-related purposes differs across racial and ethnic groups.^38^ For example, older Black and Hispanic adults are less likely than their White counterparts to use technology for health-related activities, even after accounting for demographic characteristics, education and health conditions.^38^ Likewise, our study deploys a single native application (Beiwe), predominantly on smartphone devices.^2^ We hope that readers will note that our emphasis on data missingness is not specific to any particular population or digital device. We hope that future work will apply our framework and reporting more broadly, to a more diverse array of digital devices and populations.

Overall, we hold a microscope to our own digital phenotyping data. Our results suggest that data missingness is not simply a statistical anomaly to be mitigated through imputation, but rather, a feature of possible clinical relevance. Accordingly, we suggest that data missingness should be accounted for and clearly reported in research studies at multiple timescales (day, hour, and minute), depending on the clinical context. We also propose a framework and nomenclature for the “mechanisms” of data missingness that relate directly to the clinical context of data collection. We further describe and detail the logic of a method by which clinical inference might be adjusted according to the extent of missing data, to be evaluated on a study-by-study basis.

## Data Availability

All data produced in the present study are available upon reasonable request to the authors after publication.

## Acknowledgements

The author(s) declare financial support was received for the research, authorship, and/or publication of this article. We acknowledge and are grateful for support from the National Institute on Aging (1K01AG078127-01, PI: DSB) and the Brain & Behavior Research Foundation (29966, PI: DSB).

---

**Supplementary Figure 1.**
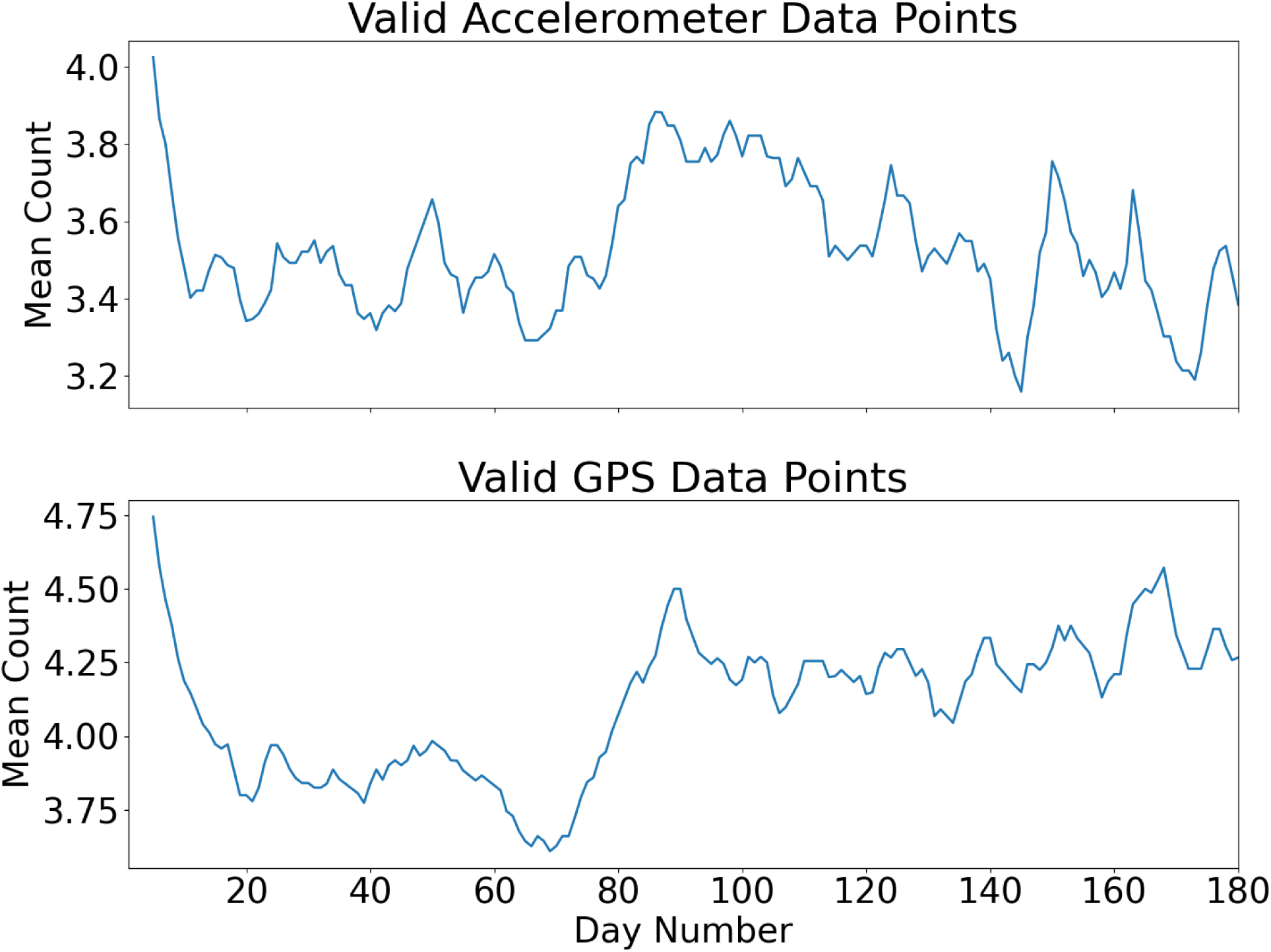
Mean count of valid data points across participants throughout the study period. This figure shows the rolling mean counts of valid data points for accelerometer (top) and GPS (bottom) measures across participants on each day. The count was calculated using a sliding window of 5 days, requiring all 5 days to have data present. The mean counts remain relatively stable and high over time, indicating a consistent level of data contribution from participants throughout the 180-day study period.

**Supplementary Figure 2.**
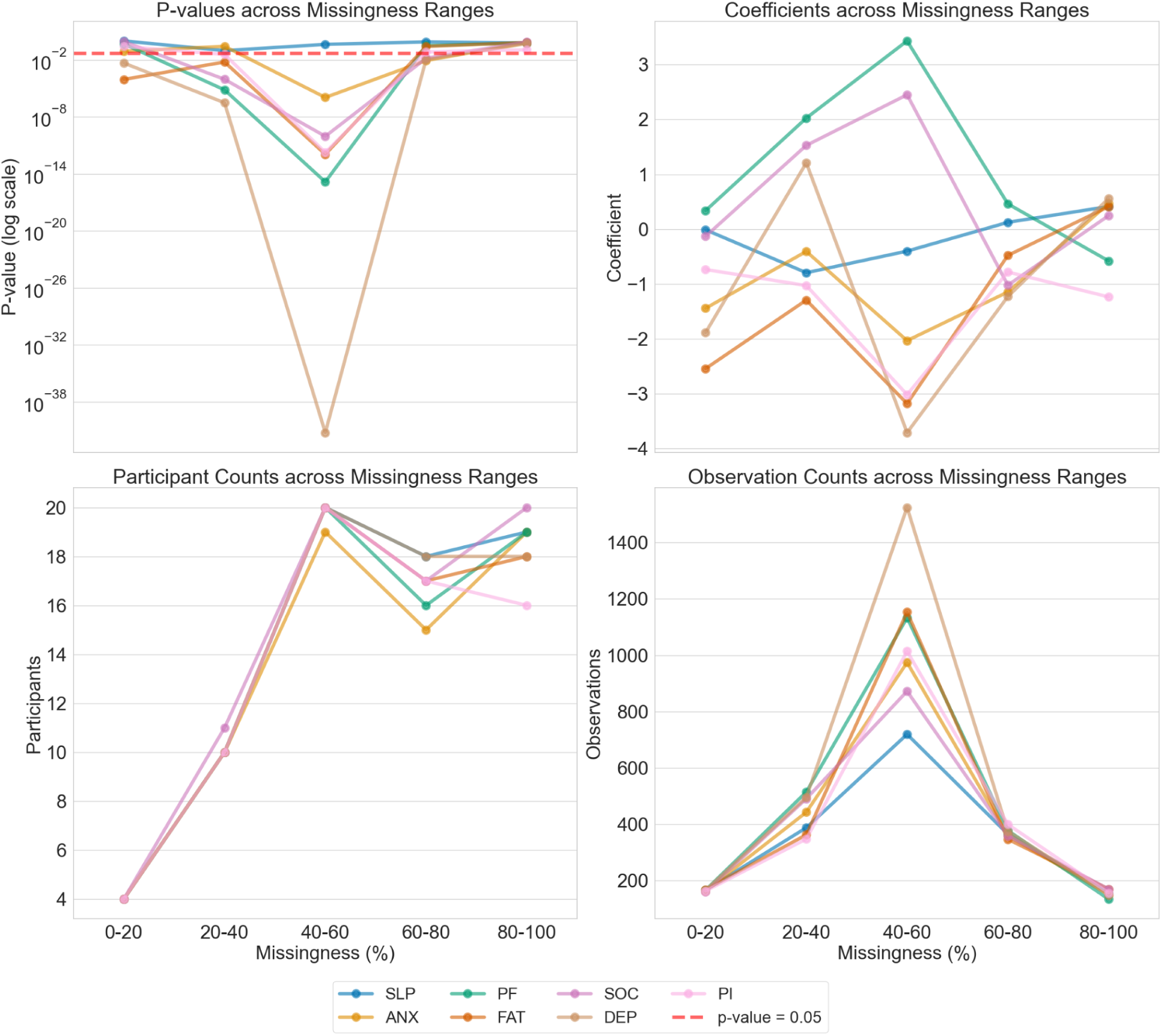
OLS analysis of cadence across hour-level data missingness ranges. This figure shows p-values (log scale), coefficients, participant counts, and observation counts across different missingness ranges (0-20%, 20-40%, 40-60%, 60-80%, 80-100%) for cadence. The p-values, participant counts, and observation counts follow similar changing patterns, albeit to different degrees, across PROMIS categories, while coefficients exhibit greater variability. The 40-60% missingness bracket stands out as a turning point, particularly for p-values and observation counts, where significant changes in the models start to occur. Each line represents a different PROMIS category, highlighting the relationships and trends as missingness increases.

**Supplementary Figure 3.**
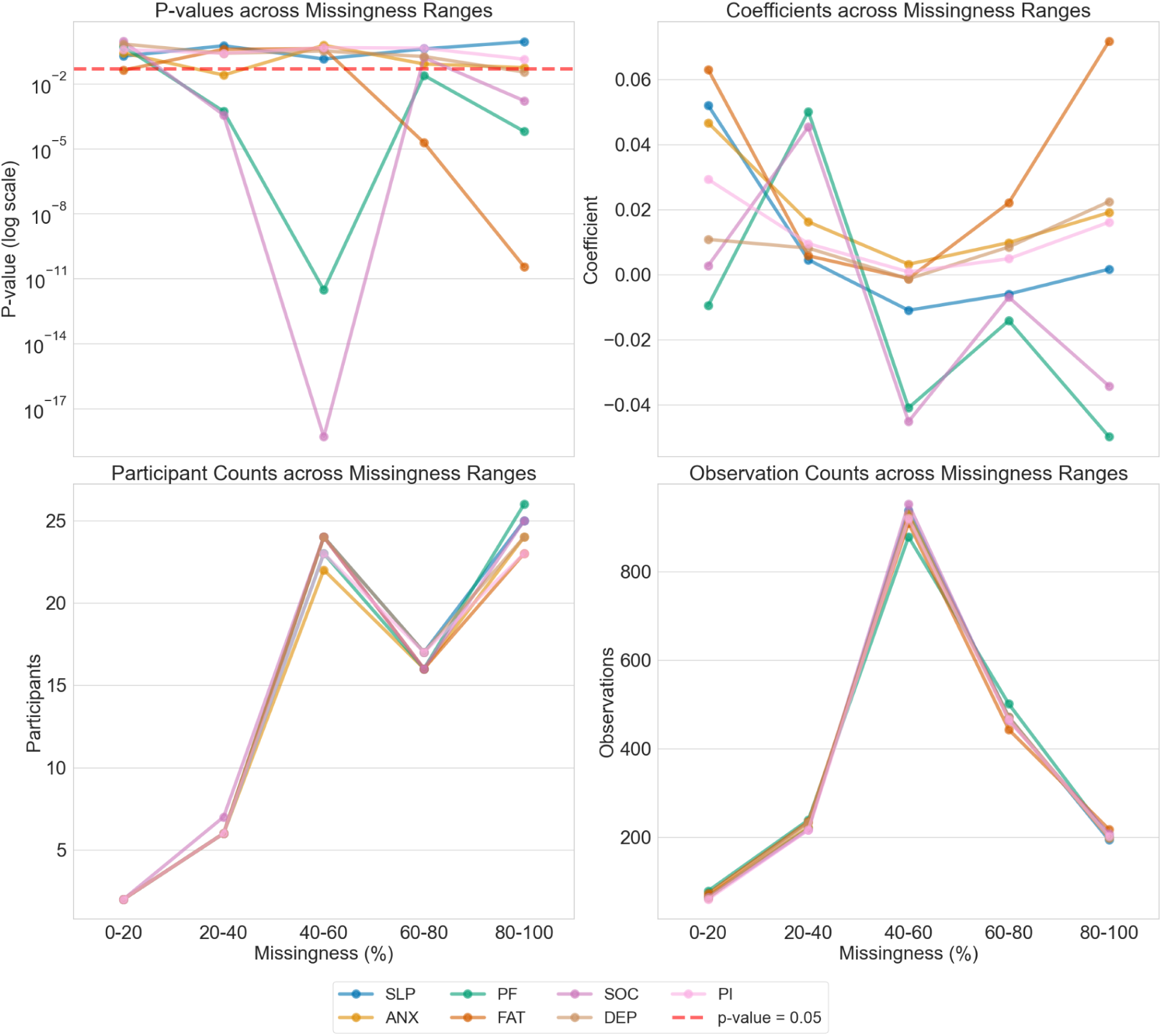
OLS analysis of home time across hour-level data missingness ranges. This figure shows p-values (log scale), coefficients, participant counts, and observation counts across varying missingness ranges (0-20%, 20-40%, 40-60%, 60-80%, 80-100%) for home time. While the p-values and coefficients exhibit varying changing patterns across PROMIS categories, the participant and observation counts remain very similar. The 40-60% missingness bracket is a turning point, particularly for observation counts, where significant changes in the models start to occur. Each line represents a different PROMIS category, highlighting the relationships and trends as missingness increases.

